# Digitized smart surveillance in malaria elimination programme in Mangaluru city, Karnataka, India ― a detailed account of operationalization in post-digitization years

**DOI:** 10.1101/2020.07.06.20147405

**Authors:** B. Shantharam Baliga, Shrikala Baliga, Animesh Jain, Naveen Kulal, Manu Kumar, Naren Koduvattat, B. G. Prakash Kumar, Arun Kumar, Susanta K. Ghosh

## Abstract

**Background:** An indigenously developed digital handheld Android-based geographical information system (GIS)-tagged tablets (TABs) device has been deployed in Mangaluru city, Karnataka, India for smart surveillance in malaria elimination programme from October 2014. Here a detailed account is enumerated in the post-digitization years. The study was aimed to assess the effectiveness of the digitized surveillance system under the ongoing health system in Mangaluru city.

**Methods:** A software developed for digitization of malaria surveillance was continued in the post-digitization years (PDY). The same digitization year (DY) protocol was followed in the post-digitization periods also. Secondary data from the malaria control software, total nunber of cases, active surveillance, malaria indices, and feedback from stakeholders were looked at and analyzed.

**Results:** Digital surveillance was sustained and the performance improved in the 5^th^ year with participation of all stakeholders. Malaria indices significantly reduced to about 65% in the digitization years compared with digitization year (*p*<0.001). Slide positivity rate (SPR) decreased from 10.36 (DY) to 4.3 (PDY4). Annual parasite incidence (API) decreased from 16.17 (DY) to 5.4 (PDY4). There was a tempo-spatial correlation between closure of cases on 14^th^ day and incidence of malaria. There was a negative correlation between contact smears and incidence of malaria (*r* = -0.907). Good impact was recorded in the pre-monsoon months (∼85%) and low impact in July and August months (∼40%).

**Conclusion:** Software helped to improve incidence-centric active surveillance, complete treatment with documentation of elimination of parasite, targeted vector control measures. The learnings and analytical output from the data helped to modify strategies for local control of both disease and the vector.

## Background

Mangaluru is a coastal city in Karnataka of southwestern India. The city is also notoriously known for being endemic for malaria for three decades [1-3]. Smart surveillance of malaria cases was introduced as programme management tool in this city in 2015. In a previous report, a detailed information was described on the design and implementation of this digitization protocol and presented initial secondary data analyzes to determine the impacts in the 2^nd^ years post-digitization [4]. Subsequent to digitization of entire malaria control programme in the territory of the city corporation, there has been a reduction in the incidence of malaria.

The current operational functionalities were assessed in the post-digitization periods, and attempted to know whether any kind of fatigues or changes in priorities had crept in the surveillance system; or any further improvements are needed to make the device more user-friendly. All malariometric indices and interventions on vector management parameters were recorded and analyzed. Routine monitoring and strict vigils were put in place on the ongoing newly introduced surveillance system using geographical information system (GIS)-tagged tablets (TABs). This paper presents the latest data, and discuss how the surveillance continued using impact data after 4-year post-digitization. It also ascertained the lessons learnt, possible analyses, hypotheses, and explanations regarding the utility of the software for surveillance, and the experiences during implementation process.

## Methods

Digitization of surveillance was initiated from October 2014 to September 2015 considering digitization year (DY) [4]. Similarly, from October 2015 to September 2016 was post-digitization year 1 (PDY 1); from October 2016 to September 2017 post-digitization year 2 (PDY 2); from October 2016 to September 2018 post-digitization year 3 (PDY 3) and from October 2018 to September 2019 post-digitization year 4 (PDY 4). Hence, malaria control software is being used for the 5^th^ consecutive year and cases are reported by all the health care providers and stakeholders. Field activities for control and closure of cases/source elimination of mosquito breeding sites were carried out based on the inputs into the software. This data available on the system was translated into excel sheet and the data was analyzed for taking appropriate decisions and amendments in action plan.

Mangaluru city has administrative units designated as wards, and 60 such wards constitute city limits [5,6]. Malaria indices were analyzed in each month in all these wards covering the entire city limits. Based on the data, high risk areas were identified periodically to carry out necessary anti-malarial activities.

Malaria cases reported in the city were also analyzed based on the type of health facilities from where patients sought health care services. These health care facilities were categorized as private health facilities, and public health facilities. Private health facilities included all the hospitals, nursing homes and diagnostic laboratories. Public health facilities included surveillance team of district vector borne disease control office (DVBDCO), government-run hospitals, urban health centres, and malaria clinics.

Each incidence was analyzed based on reporting time, closure of the cases, closure within 14 days after complete treatment and follow-up smear examination for persistence of parasites. Closure time is considered as 14 days to complete primaquine therapy for *Plasmodium vivax* cases to prevent relapse as per the recommendation of National Vector Borne Disease Control Programme (NVBDCP) [7].

Factual reporting with regards to administrative decisions, hurdles in the implementation of anti-malarial activities, and how these problems were addressed and their effects on the malaria control and the malaria indices were analyzed.

### Statistical analysis

Closing of each positive case and also vector interventions were analyzsed following the scattered plot method. Community visits, contact smears during active surveillance around reported case (ASARC), vector control activities were analyzed along with malaria indices such as Annual Blood Examination Rate (ABER), Slide Positivity Rate (SPR), Slide Falciparum Rate (SFR) and Annual Parasite Incidence (API). Mothly malaria trends at each level were also plotted in relation to closure of cases. Impact on these parameters were also analysed using Chi-square tests and Pearson’s correlation coefficient formula were applied wherever appropriate using IBM SPSS Statistics 25. Significance values *p*<0.001 was considered significant.

## Results

Monthly malria incidence for the past 6 years and the cumulative reduction in incidence in the urban limits of Mangalore is depicted in Table 1. The gradual reduction of overall incidence of malaria continued in the 4^th^ year post-digitization (PDY4) with an overall cumulative reduction by 65% as compared to the digitization year (DY).

**Table 1:** Monthly incidence of malaria*.

| Month | 2013- 14 | 2014-15<br>DY | 2015-16<br>PDY1 # | 2016-17<br>PDY2 | 2017-18<br>PDY3 | 2018-19<br>PDY4 | Cumulative<br>reduction<br>(%) (PDY4) | 2019-20<br>PDY5 |
| --- | --- | --- | --- | --- | --- | --- | --- | --- |
| October | 479 | **1064** | 929 | 717 | 776 | **398** | 62.5 | 357 |
| November | 465 | **1278** | 1116 | 631 | 750 | **458** | 64.1 | 190 |
| December | 454 | **1103** | 1348 | 468 | 728 | **412** | 62.6 | 209 |
| January | 532 | **1101** | 1068 | 403 | 438 | **342** | 68.9 | 145 |
| February | 471 | **554** | 662 | 305 | 281 | **182** | 67.1 | **87** |
| March | 477 | **521** | 400 | 305 | 329 | **180** | 65.4 | **54** |
| April | 617 | **528** | 475 | 405 | 294 | **117** | 77.8 | **79** |
| May | 1004 | **715** | 449 | 374 | 384 | **106** | 85 | **64** |
| June | 1159 | **1065** | 1142 | 656 | 741 | **166** | 84 | - |
| July | 1526 | **971** | 2084 | 1174 | 1003 | **581** | 40 | - |
| August | 1062 | **848** | 1952 | 1325 | 837 | **514** | 41 | - |
| September | 1421 | **1224** | 989 | 874 | 549 | **294** | 75 | - |
| **Total** | **9667** | **10972** | **12641** | **7637** | **7110** | **3750** | 65 | - |

*\*malaria incidence decreased to double digit in the 5<sup>th</sup> year; the data is current as on date of submission of manuscript.*
*# some diagnostic centres reported cases directly to malria control cell*

The cumulative reduction in the incidence was also calculated (Table 2). The maximum reduction of 84 to 85% was noted for the months of May and June and least 40 to 41% in the months of July and August. Overall reduction in incidence was remarkable at 65%. The trend has continued and malaria cases decreased to double digits consistently for 4 months in the 5^th^ year of smart surveillance.

**Table 2:** Ward-level malaria cases in Mangalore post-digitization and cumulative reduction.

| Ward | PDY1 | PDY2 | PDY3 | PDY4 | Cumulative reduction (%) |
| --- | --- | --- | --- | --- | --- |
| 1-Surathkal - West | 105 | 89 | 18 | 26 | -75 |
| 2-Surathkal - East | 55 | 40 | 13 | 22 | -60 |
| 3-Katipalla- East | 36 | 13 | 11 | 5 | -86 |
| 4-Katipalla-K'pura | 17 | 16 | 13 | 11 | -35 |
| 5-Katipalla - North | 22 | 10 | 14 | 10 | -55 |
| 6-Idya – East | 49 | 126 | 68 | 26 | -47 |
| 7-Idya – West | 25 | 15 | 17 | 7 | -72 |
| 8-Hosabettu | 18 | 23 | 17 | 12 | -33 |
| 9-Kulai | 0 | 20 | 32 | 8 | -60 |
| 10-Baikampady | 77 | 44 | 40 | 22 | -71 |
| 11-Panambur | 101 | 69 | 50 | 55 | -46 |
| 12-Panjimogaru | 91 | 51 | 53 | 33 | -64 |
| 13-Kunjathbail -North | 77 | 33 | 51 | 24 | -69 |
| 14-Marakada | 56 | 17 | 54 | 28 | -50 |
| 15-Kunjathbail- south | 60 | 37 | 61 | 20 | -67 |
| 16-Bangrakulur | 74 | 73 | 88 | 52 | -30 |
| 17-Derebail - North | 459 | 128 | 207 | 61 | -87 |
| 18-Kavoor | 397 | 105 | 115 | 72 | -82 |
| 19-Pachanady | 101 | 41 | 18 | 20 | -80 |
| 20-Thiruvail | 31 | 14 | 0 | 8 | -74 |
| 21-Padavu - West | 66 | 32 | 82 | 28 | -58 |
| 22-Kadri Padavu | 291 | 209 | 135 | 82 | -71 |
| 23-Derebail - East | 432 | 224 | 105 | 120 | -72 |
| 24-Derebail - South | 109 | 254 | 277 | 51 | -53 |
| 25-Derebail - West | 246 | 157 | 293 | 166 | -33 |
| 26-Derebail - N-East | 212 | 123 | 201 | 159 | -25 |
| 27-Bolloor | 176 | 39 | 45 | 26 | -85 |
| 28-Mannagudda | 346 | 110 | 170 | 31 | -91 |
| 29-Kambla | 85 | 90 | 46 | 14 | -84 |
| 30-Kodialbail | 186 | 134 | 140 | 65 | -65 |
| 31-Bejai | 162 | 157 | 236 | 132 | -19 |
| 32-Kadri – North | 131 | 75 | 55 | 107 | -18 |
| 33-Kadri – South | 181 | 159 | 341 | 211 | +17 |
| 34-Shivabagh | 113 | 74 | 93 | 47 | -58 |
| 35-Padavu - Central | 120 | 124 | 124 | 56 | -53 |
| 36-Padavu – East | 121 | 107 | 118 | 94 | -22 |
| 37-Maroli | 107 | 65 | 58 | 15 | -86 |
| 38-Bendoor | 205 | 113 | 74 | 29 | -86 |
| 39-Falnir | 240 | 92 | 55 | 21 | -91 |
| 40-Court | 293 | 322 | 438 | 180 | -39 |
| 41-Central Market | 710 | 644 | 404 | 191 | -73 |
| 42-Dongarkeri | 104 | 66 | 40 | 39 | -63 |
| 43-Kudroli | 218 | 58 | 65 | 31 | -86 |
| 44-Bunder | 799 | 496 | 400 | 248 | -69 |
| 45-Port | 753 | 554 | 451 | 222 | -71 |
| 46-Cantonment | 147 | 227 | 292 | 156 | + 6 |
| 47-Milagrese | 507 | 233 | 163 | 69 | -86 |
| 48-Kankanady<br>Valencia | 370 | 133 | 95 | 22 | -94 |
| 49-Kankanady | 255 | 72 | 70 | 29 | -89 |
| 50-Alape – South | 108 | 50 | 37 | 21 | -81 |
| 51-Alape – North | 109 | 84 | 32 | 10 | -91 |
| 52-Kannur | 43 | 29 | 36 | 18 | -58 |
| 53-Bajal | 118 | 48 | 30 | 8 | -93 |
| 54-Jeppinamogaru | 62 | 41 | 27 | 12 | -81 |
| 55-Attavara | 307 | 181 | 101 | 54 | -82 |
| 56-Mangaladevi | 418 | 174 | 181 | 64 | -85 |
| 57-Hoige Bazaar | 278 | 412 | 205 | 112 | -60 |
| 58-Bolar | 197 | 109 | 78 | 49 | -75 |
| 59-Jeppu | 132 | 79 | 59 | 24 | -82 |
| 60-Bengre | 446 | 323 | 294 | 193 | -55 |

In June 2018, malaria elimination teams were formed to visit reported cases of malaria and carry out sanitization of the area subsequent to administrative decision to utilize services of multipurpose workers (MPWs) for non-malarial work. The resultant figures for incidence in PDY 2018-19 show that there were marked reductions in malaria cases.

Ward-level malaria incidence was also recored in each 60 ward shown in Table 2. The ward-level cumulative reduction in incidence of malaria from the PDY1 to PDY4 was significant (*p*<0.001). However, two wards (Kadri South and Cantonment) showed increase in incidence, with one of the wards showing 17% increase in the cases compared to the baseline value of PDY1.

It was noted that surveillance continued to improve with malaria cases being reported from all the hospitals and diagnostic centres of private as well as public health system. Before digitization, private healthcare facilities contributed to nearly two thirds (68%) of the total cases being reported while the public health system contributed to nearly one third (which included 18.6% by community public hospitals and 4.3% by malaria clinics). In the post-digitization phase the contribution from private hospitals to total number of cases kept steadily declining and reduced to 57% in the 4^th^ year. At the same time, the public health system, i.e., public hospitals, urban health centres as well as DVBDCO started contributing larger proportion of total number of cases. ASARC contributed to over 1% of malaria incidence emphasizing the role played by it (Table 3).

**Table 3:** Type of health facility and malarial case reports.

|  | PDY1* | PDY2 | PDY3 | PDY4 |
| --- | --- | --- | --- | --- |
| Total cases | 11757 | 7637 | 7110 | 3750 |
| District Vector borne Disease Control Office (DVBDCO) | 571 (4.9%) | 648 (8.5%) | 593 (8.3%) | 381 (10.2%) |
| Public Hospitals | 2184 (18.6) | 1157 (15.1%) | 1406 (19.8%) | 778 (20.7%) |
| Urban health centres | 329 (2.8%) | 601 (7.9%) | 811 (11.4%) | 322 ( 8.9%) |
| Active surveillance | 123 (1.1%) | 32 (0.4%) | 55 (0.8%) | 44 (1.2%) |
| Malaria clinics | 501 (4.3%) | 327 (4.3%) | 255 (3.58%) | 89 (2.4%) |
| Private Health facilities | 8049 (68%) | 4872(63%) | 4245(56%) | 2136 (57%) |

*\*cases directly reported to malaria control cell are not included*

Table 4 depicts the number of cases for the last 4 years wherein visits were made, contact smears taken, source documented, and closure of each case was carried out. Timely field activity for vector control, complete treatment, smear to ensure parasite clearance after treatment followed by closure of cases at the earliest improved post digitization. The early reporting of cases as well as closure of cases within 14 days increased steadily.There was a negative corrrelation between the ratio of contact smears to total no. of cases and no. of positive cases detected by contact smears though it was not statistically significant.

**Table 4:**
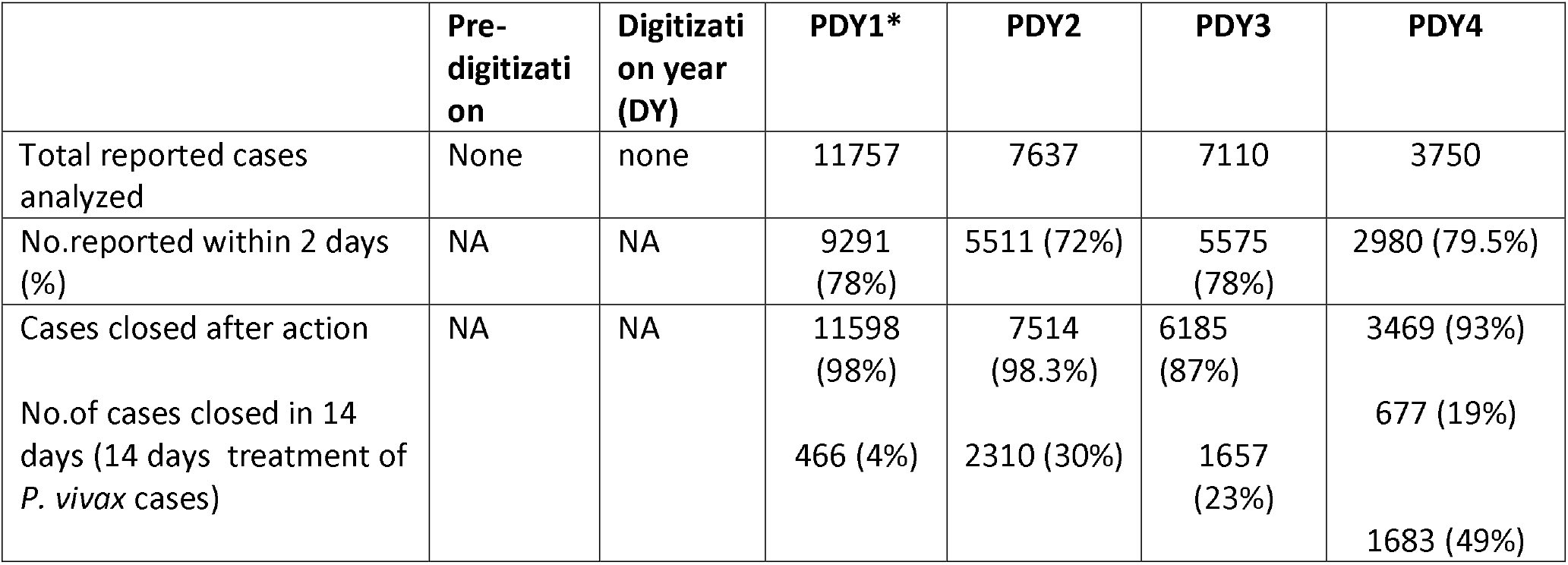
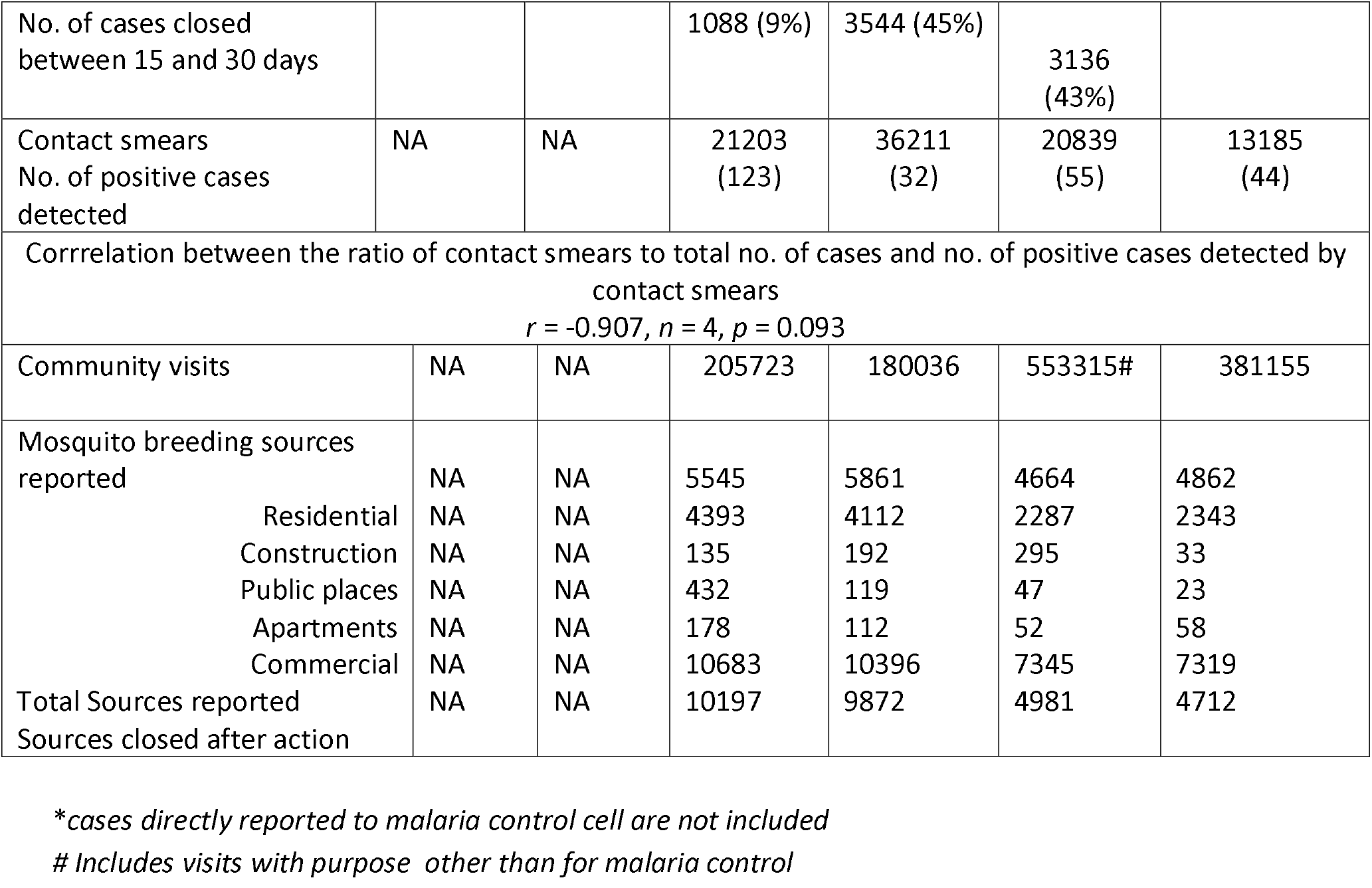
Reporting pattern, case management, smear collection source management and malarial indices before and after malaria control software introduction.

Malaria incidence and its impact on the incidence of malaria was analyzed using Pearson’s correlation coefficient formula.The ward-level closure of cases was plotted against incidence of malaria cases and correlation equation was generated as depicted in Fig. 1. The scatter plot depicts clustering of cases. It is observed that as the percentage of closure increases, the reduction in incidence is higher.The percentage of closure rate <14 days after treatment and the incidence rate of Malaria in Mangalore is depicted in Fig. 2. Each Dot represents percentage of cases treated, investigated completely and closed within stipulated period of 14 days (*r* = 0.359). As seen, the cumulative reduction of malarial cases at the end of 4 years after introduction of digitization using malaria control software (MCS) stood at 65% as compared to pre-digitization year (2014-15). The API, SPR, and SFR showed statistically significant change (*p*<0.001). The ward-level depiction based on API is shown in Fig 3. It can be noted that the wards with API in the red zone (API > 10) have reduced and the ones with green (API ≤ 2) as well as yellow (API > 2.1 to 5) have increased.

**Figure 1:**
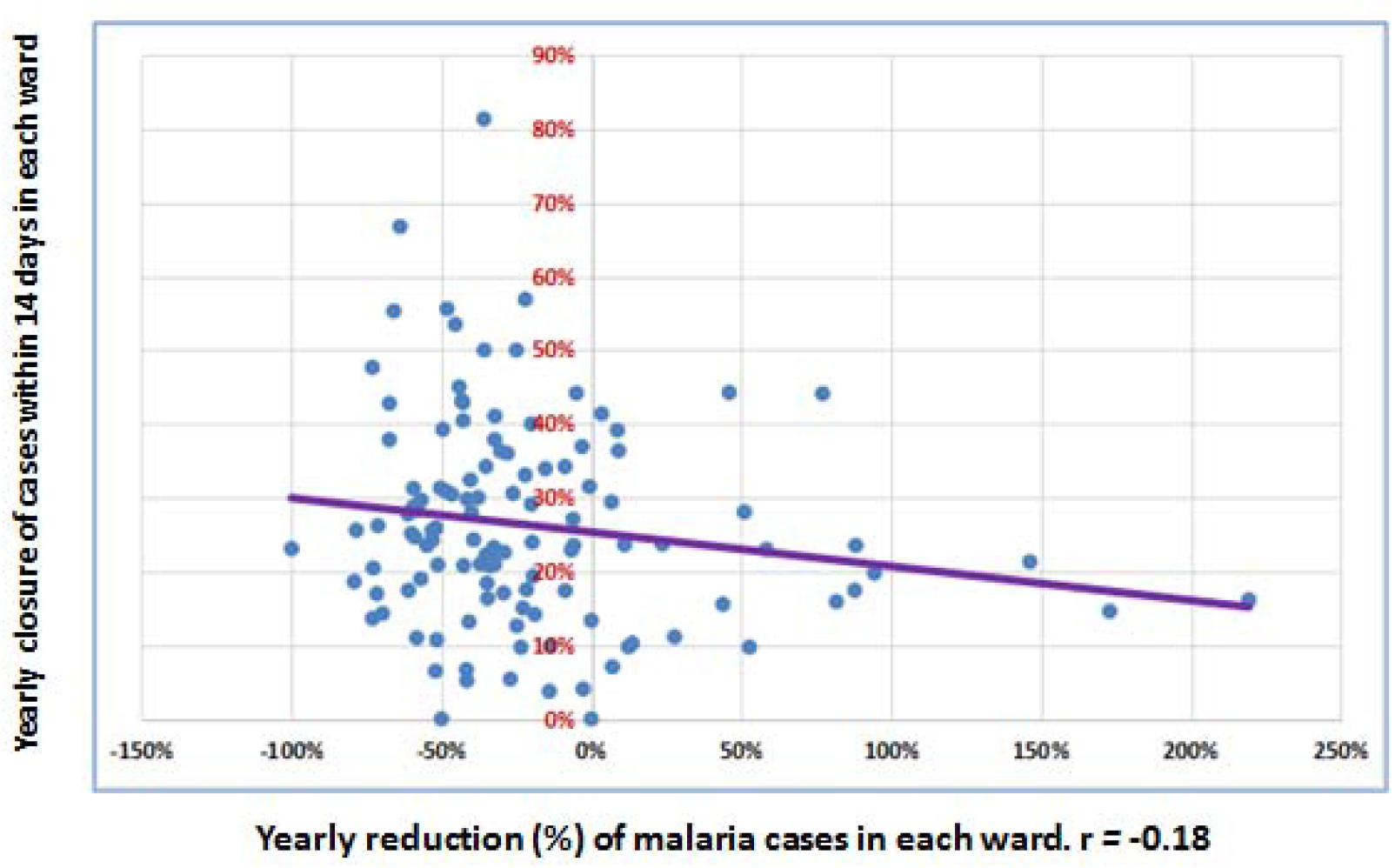
Percentage of closure rate <14 days *vs* incidence rate - trends for 4 years - administrative block (ward)-level analysis.

**Figure 2:**
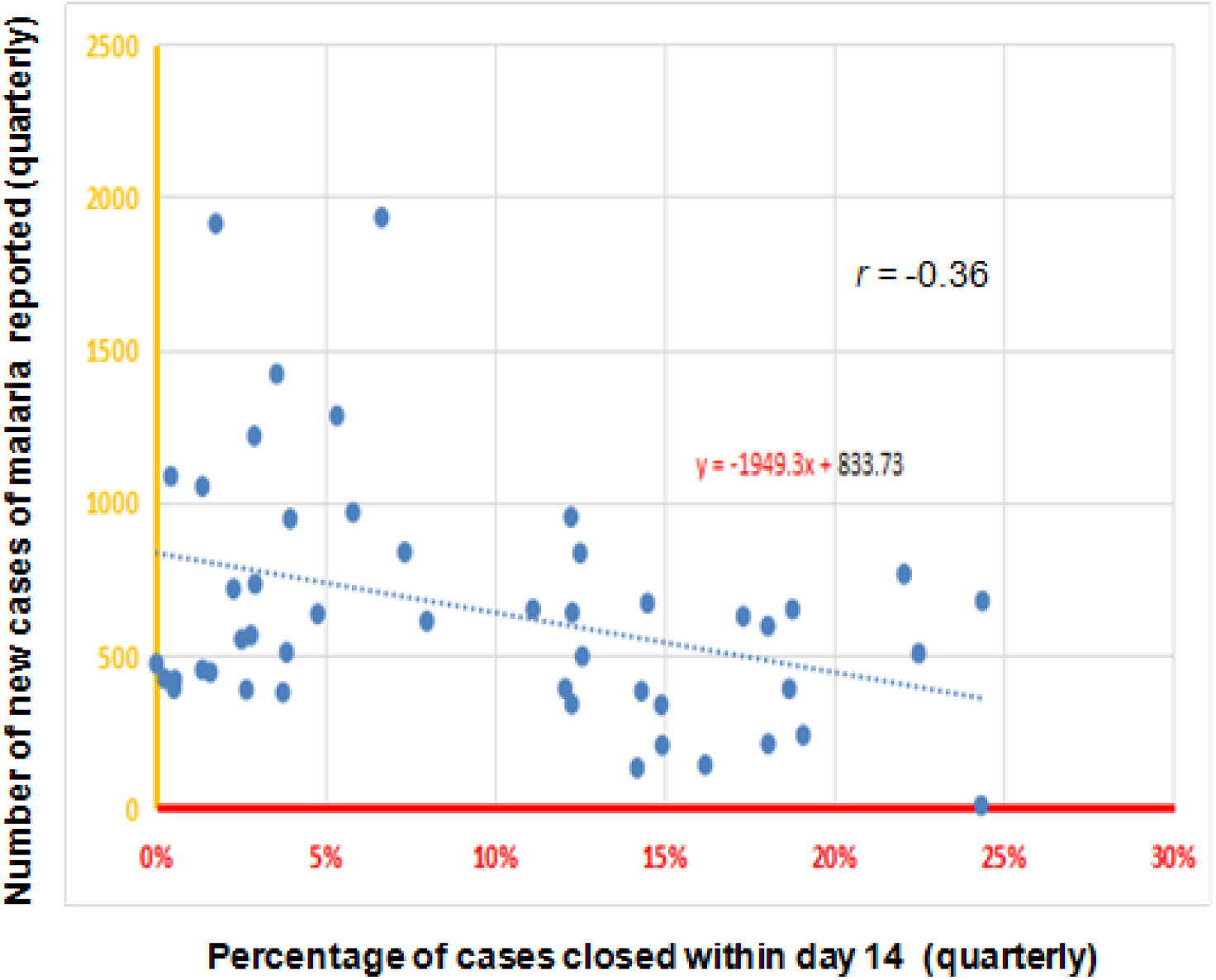
Percentage of closure rate <14 days after treatment *vs* incidence rate (quarterly) of malaria in Mangalore.

**Figure 3.**
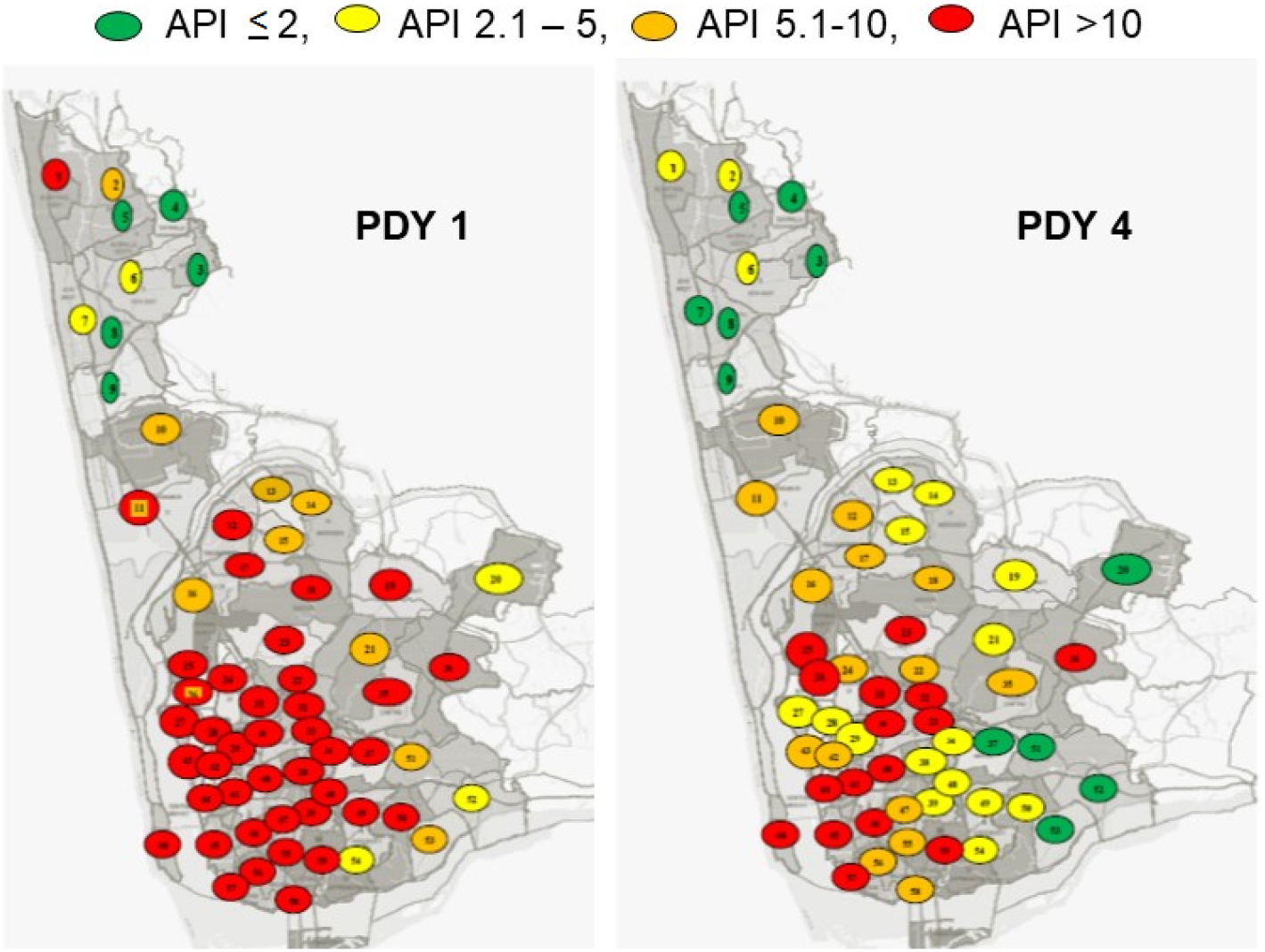
Map of Mangalore with various wards depicting the areas based on API in PDY1 and PDY4.

The malarial indices were calculated for the pre-digitization year, digitization year and each of the four years post digitization (Table 5). The slide positivity rate was seen steadily decreasing and the average annual parasite incidence (API) reduced and came down to 5.4 in the 4^th^ year post digitization.

**Table 5.** Malaria indices of Mangalore pre-digitization, digitization and post digitization year.

|  | **Pre-digitization** | **Digitization year** | **1<sup>st</sup> yr Post digitization** | **2<sup>nd</sup> yr Post digitization** | **3<sup>rd</sup> yr Post digitization\*** | **4<sup>th</sup> year Post digitization** |
| --- | --- | --- | --- | --- | --- | --- |
| Total malarial cases (no) | 8867 | 10962 | 12614 | 7637 | 7110 | 3750 |
| Number of smears/ incidence | 9.48 | 9.75 | 12.24 | 26.68 | 18.37 | 23.18 |
| Vivax malaria (% of total) | 8092 (91) | 10196(93) | 11277 (89) | 6245 (82) | 5633(79) | 3099 (82) |
| Falciparum malaria (% of total) | 775 (9) | 766(7) | 1337 ( 11) | 1395 (18) | 1494 (21) | 651 (18) |
| **Chi-square for trend $\chi^2 = 164.5$ $p$ value <0.001** |  |  |  |  |  |  |
| ABER | 13.48 | 17.13 | 24.75 | 32.68 | 20.9 | 17.75 |
| SPR | 11.15 | 10.36 | 8.17 | 3.74 | 5.4 | 4.3 |
| SFR | 0.92 | 0.73 | 0.86 | 0.68 | 1.1 | 0.7 |
| API | 15.51 | 16.17 | 18.42 | 12.24 | 11.4 | 5.4 |

*\*MPW's were given additional work other than malaria surveillance*

Inputs from stakeholders was taken regarding the challenges faced by them and how MCS was helpful in overcoming those challenges. Table 6 summarizes the challenges, gapsand functionality of the software as described by the administrator of malaria control programme in the civic body.

**Table 6.**
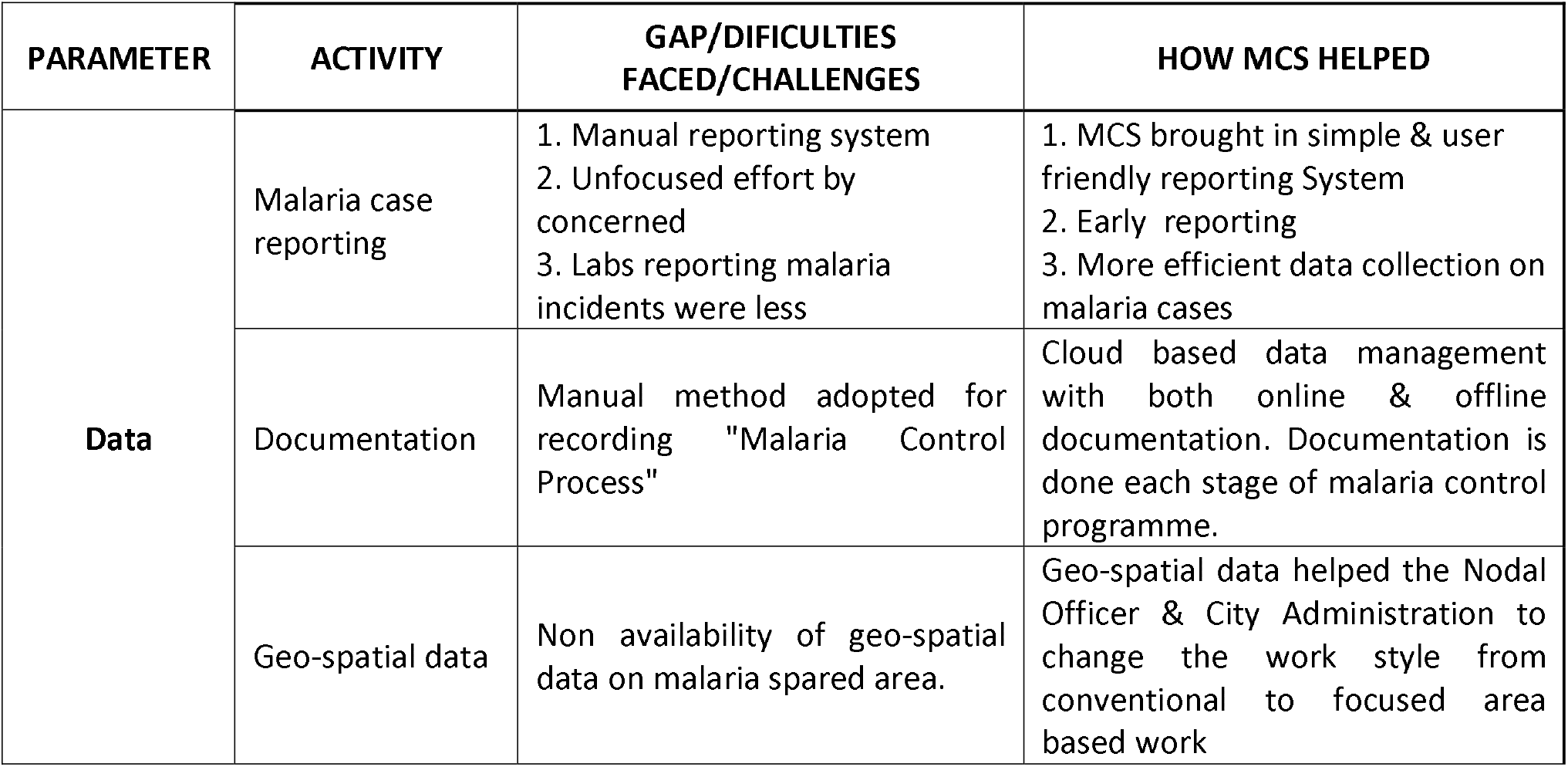
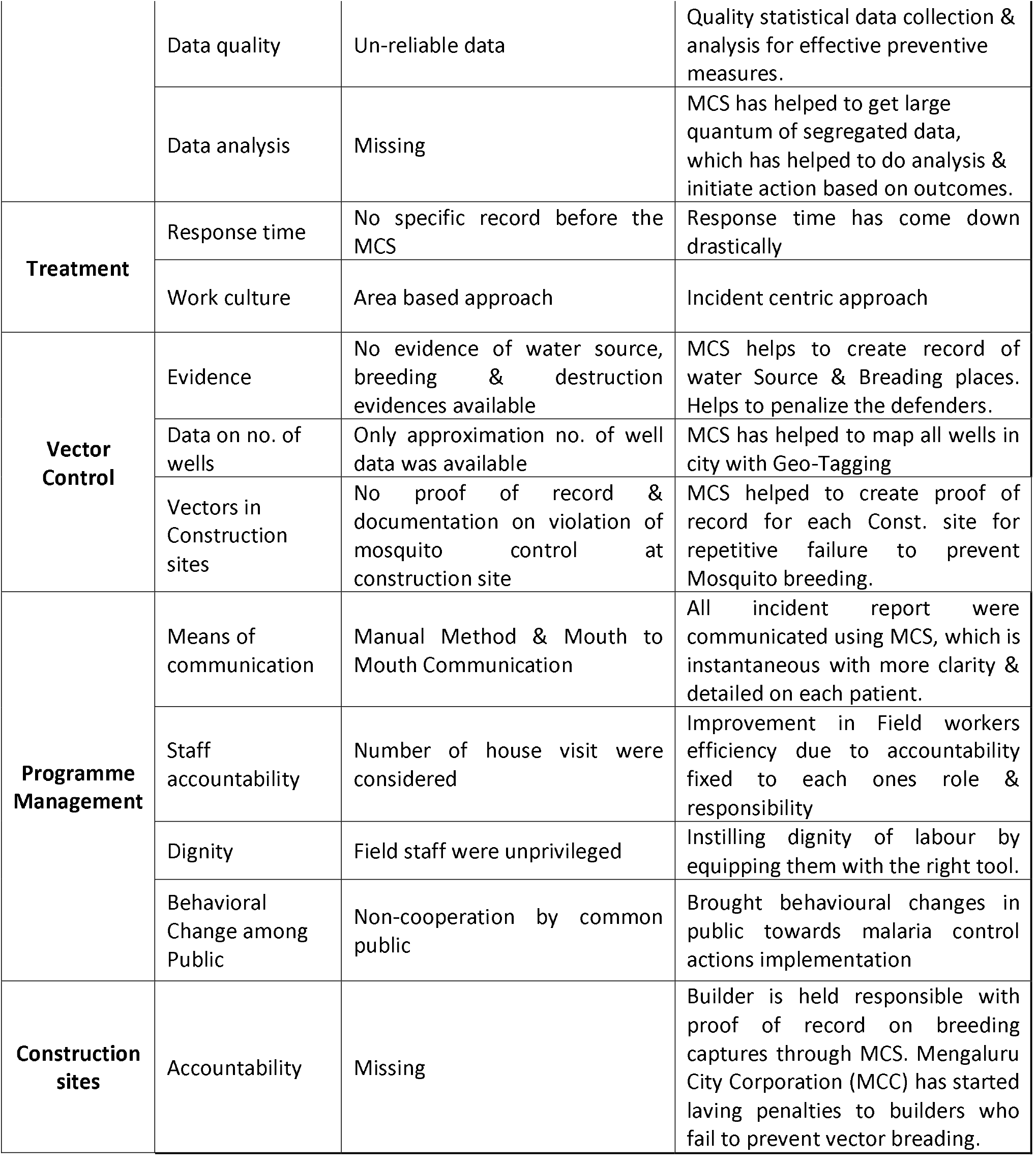
Feedback from stakeholders regarding various issues in malaria control prior to introduction of malaria control software (MCS) and description of how the software helped administrators based on inputs from stakeholders.

## Discussion

While the preliminary results were encouraging [4], it was very important to assess the impact of interventions in order to determine its future course and also to look for additional measures to strengthen the ongoing malaria control operations.The purpose of improving the reporting time and providing the TABs to MPWs was to improve surveillance and make it “smart.” Smart surveillance reduces the time delay between diagnosis and registration of a new incidence for necessary action in the field to break the chain of transmission.

There was a marked reduction in malaria incidence in Mangaluru over 4 years, and also in 2020 (up to May). It is encouraging to note that the data records show cumulative reduction by 65% and continues to decline (Table 1). For the first time in the past two decades malaria incidence reduced to double digits in the 5^th^ year as a consequence of improved surveillance and effective field work. The lower reduction in the months of July and August could be attributed to the monsoon rains and excessice mosquito breeding resulting in spread of malaria. After the introduction of malaria elimination team in June 2018, it was noted that the malaria incidence reduced even further from a peak of 1003 in July 2018 to just 294 at the end of PDY 4. The monthly closure *vs* incidence shows that with higher closure of cases, the incidence of malaria decreased. This indicates that smart surveillance does have an important role to play in breaking the chain of transmission.

Subsequent to smart surveillance, an important behavioral change took place among the diagnosticians at the point of diagnosis and it continued throught PDY4 wherin details of 80% of newly diagnosed cases were uploaded into the system within 48 hrs. These case records were available to field workers for sanitization and active surveillance in and around reported case (Table 3). Emphasis was laid on this process during implementation from the first year of programme and subsequently malaria elimination team was formed for rapid response. After effective implementation of control programme aided by software for 18 months, an administrative decision was taken to utilize services of MPWs for non-malarial (civic body’s) work resulting in reduced efficiency in the field. Although the community visits increased by manifold during PDY3, it was not translated to effective vector control measures and collection of smears by active surveillance reduced from 4.61 per case (PDY2) to 2.8 per case (PDY3). This resulted in slump in the work and lesser reduction of malarial incidences during PDY3. A surge in the number of cases was observed in April – May 2017 which led to increase in malarial indices. To counter this inefficiency, Complete Malaria Elimination Teams (CMETs) were formed at district malaria unit in June 2018. These teams visited each malaria case as soon as the case details were uploaded and conducted ASARC along with anti-vector activities in the locality. First visit and collection of smears from contacts and fever cases in the residences around reported case did help in brealking transmission as observed by the negative correlation between contact smear and to incidences of malaria over 4 years.

Even ward-level analysis demonstrated that the incidence in almost all the wards was reducing progressively with a good cumulative reduction in incidence(Table 2). However, it can be noted that the effect of the entire programme, its implementation and effects on malaria control was not uniform in all administrative units (wards). Most wards showed reduction ranging between 23% to 94%. The maximum reduction was noted in Kankanady-Valencia ward which had a reduction of 94% in PDY4, followed by Bajal which had a reduction of 93%. Although certain wards had low incidence, there was an increase in cases in 2 wards namely Kadri South and Cantonment. Kadri South ward showed a cumulative increase of 17% while Cantonment ward showed an increase of 7%. This is attributed to an increase in the number of cases in both these wards in PDY3 due to the repurposing of the MPWs for non-malaria work. Kadri south ward had recorded 341 cases in PDY3 compared to 159 in PDY 2 (114% increase) and Cantonment had an increasefrom 227 in PDY 2 to 292 cases in PDY 3 (29% increase). However, even these two wards do show a progressive reduction by PDY3 when the malaria elimination teams were deployed. These wards, where there was an increase in cases or very minimal reduction in cases (less than 20% in 4 years) against the expected trend as seen in other wards may probably be indicative of problematic areas. High risk categorization is based on API and such wards recorded reduction of incidence by 80% and above. Several wards converted from a high API red zone to a lesser API green or yellow zones (Fig. 3). This assumes greater significance in the light of a recent report about the asymptomatic malaria carriers in hotspots of malaria at Mangalore which indicates that these may seed transmission to the surrounding population in receptive areas [8]. There may be a role to understand geographic trends for planning the strategies at micro level and further research and review of these is warranted. Further, it may be worthwhile to look at the sociodemographic cahracteristics of people in these areas as well as the activities like construction and migration or travel [9].

Private sector contribution was higher than the public health system. According to WHO, reported cases of malaria are only from public health care facilities^10,11^ and hence, large number is unreported. However, even where reporting rates in the public health sector are close to a 100%, in some countries, more than 50% of malaria patients seek care in the private sector [12]. With digitization both public and private health care providers reported the malarial cases (Table 3). Private sector contribution was higher than the public health system. According to WHO reported cases of malaria are only from public health care facilities [10,11] and a large number is unreported. However, even where reporting rates in the public health sector are close to a 100%, in some countries, more than 50% of malaria patients seek care in the private sector [12]. Thus the software helped to connect people at point of diagnosis from both private and public health systems with field workers instantaneously for sanitization exercise, investigating contacts for malarial parasites and ensuring complete elimination of parasite from malarial patient. This was the biggest advantage of smart surveillance which is the essence of this software.

The action of closing the caseon day 14, which reflects accountability of field force steadily increased and it did contribute to reduction of malaria cases. The proportion of closure of cases between 15 – 30 day was also steadily maintained. There was both temporal and spatial relation to this action of field force (Figs 1 and 2). Greater the monthly closure of cases the higher was the decrease in incidence of malaria in a geographical area (Fig. 1). Hence, it ensures completion of treatment and ASARC establishes that breaking the chain of transmission and measures to reduce breeding and spread are important public health measures in control of malaria [4]. The software helped in this activity and also aided in monitoring the activities of MPWs and closure of cases with documentation.

Data from software was analyzed for changing the approach to field activities for malaria control. As described earlier, surveillance was carried out as soon as new malarial case was reported − ASARC. During analysis of new cases, clusters of new cases within a short period of one week, within a defined geographical area were identified and strategically separate programmes were carried out. One such endeavour was targeted for labourers/ daily wage earners. Generally, malaria clinics are open from 9 AM and to 5 PM which were underutilized as it was not convenient for the manual labourers/daily wage earners and low socioeconomic class, as they were engaged in their vocation and income generation activities during that time. Hence, a mobile 24×7 clinic using a van and health care workers was introduced so that it could visit various places and could also be sent to the site if there was a phone call made to the central malaria helpline number. This helped in not only enhancing the diagnosis but also treatment and prompt reporting of malaria cases.

Smart surveillance did empower the stake hoders. Table 7 summarises perception of stakeholders and how the software was utilized and how it brought about positive change in them.

**Table 7.**
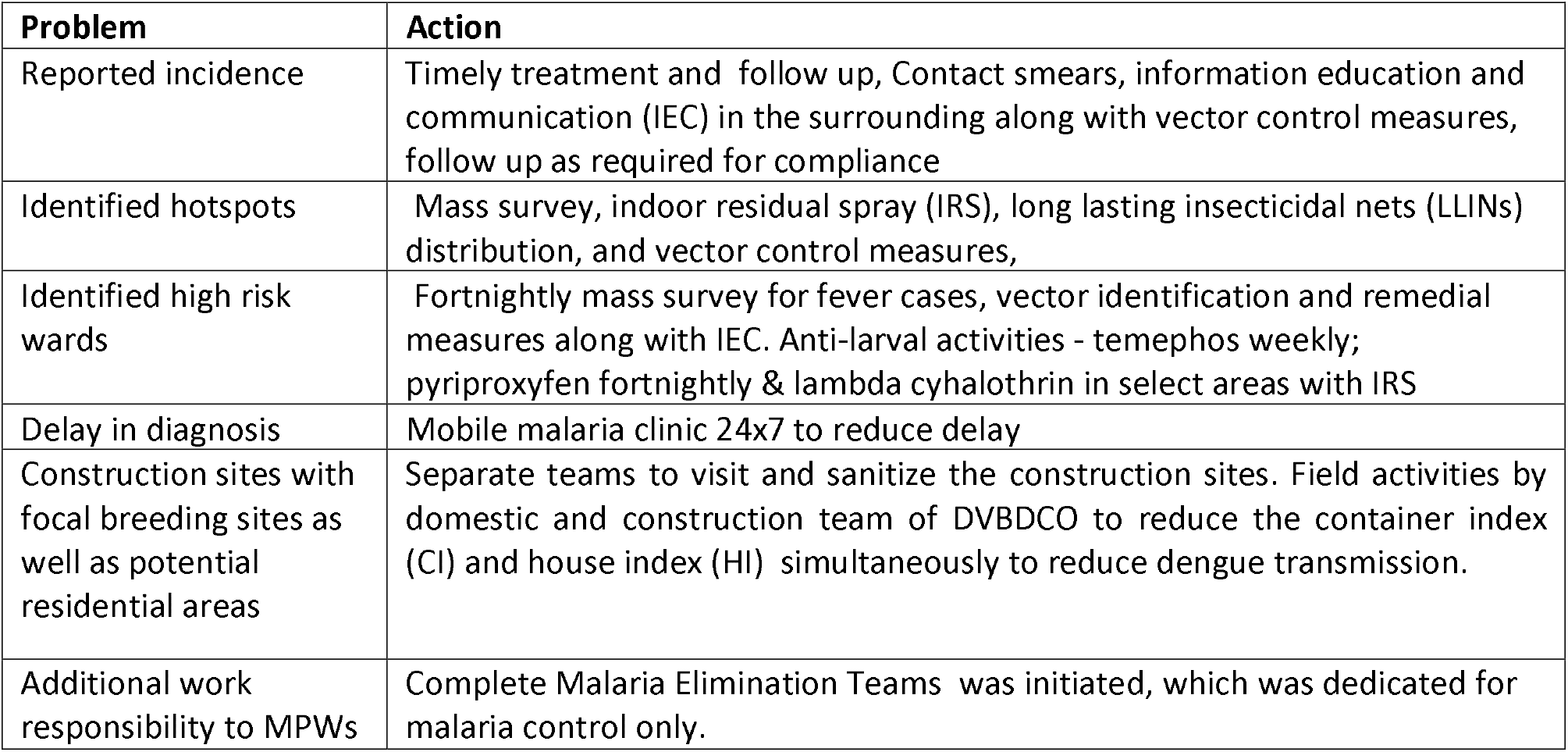
Summary of strategies adopted based on analysis of data obtained from software and addressing the issues.

| **Problem** | **Action** |
| --- | --- |
| Reported incidence | Timely treatment and follow up, Contact smears, information education and communication (IEC) in the surrounding along with vector control measures, follow up as required for compliance |
| Identified hotspots | Mass survey, indoor residual spray (IRS), long lasting insecticidal nets (LLINs) distribution, and vector control measures, |
| Identified high risk wards | Fortnightly mass survey for fever cases, vector identification and remedial measures along with IEC. Anti-larval activities - temephos weekly; pyriproxyfen fortnightly & lambda cyhalothrin in select areas with IRS |
| Delay in diagnosis | Mobile malaria clinic 24x7 to reduce delay |
| Construction sites with focal breeding sites as well as potential residential areas | Separate teams to visit and sanitize the construction sites. Field activities by domestic and construction team of DVBDCO to reduce the container index (CI) and house index (HI) simultaneously to reduce dengue transmission. |
| Additional work responsibility to MPWs | Complete Malaria Elimination Teams was initiated, which was dedicated for malaria control only. |

Fig. 4 describes how the information technology (IT) software functions to improve control activity. All the actions regarding treatment and vector control can be measured including the time frames for each activity. Transmission cycle is effectively broken if interventions are carried out in the first 10 days of diagnosis in addition to early diagnosis. Transmission occurs locally around a reported case and it is logical to implement effective vector control and measure that activity. Smart surveillance was able to measure many control measures which helped to change the strategies in the field.

**Figure 4:**
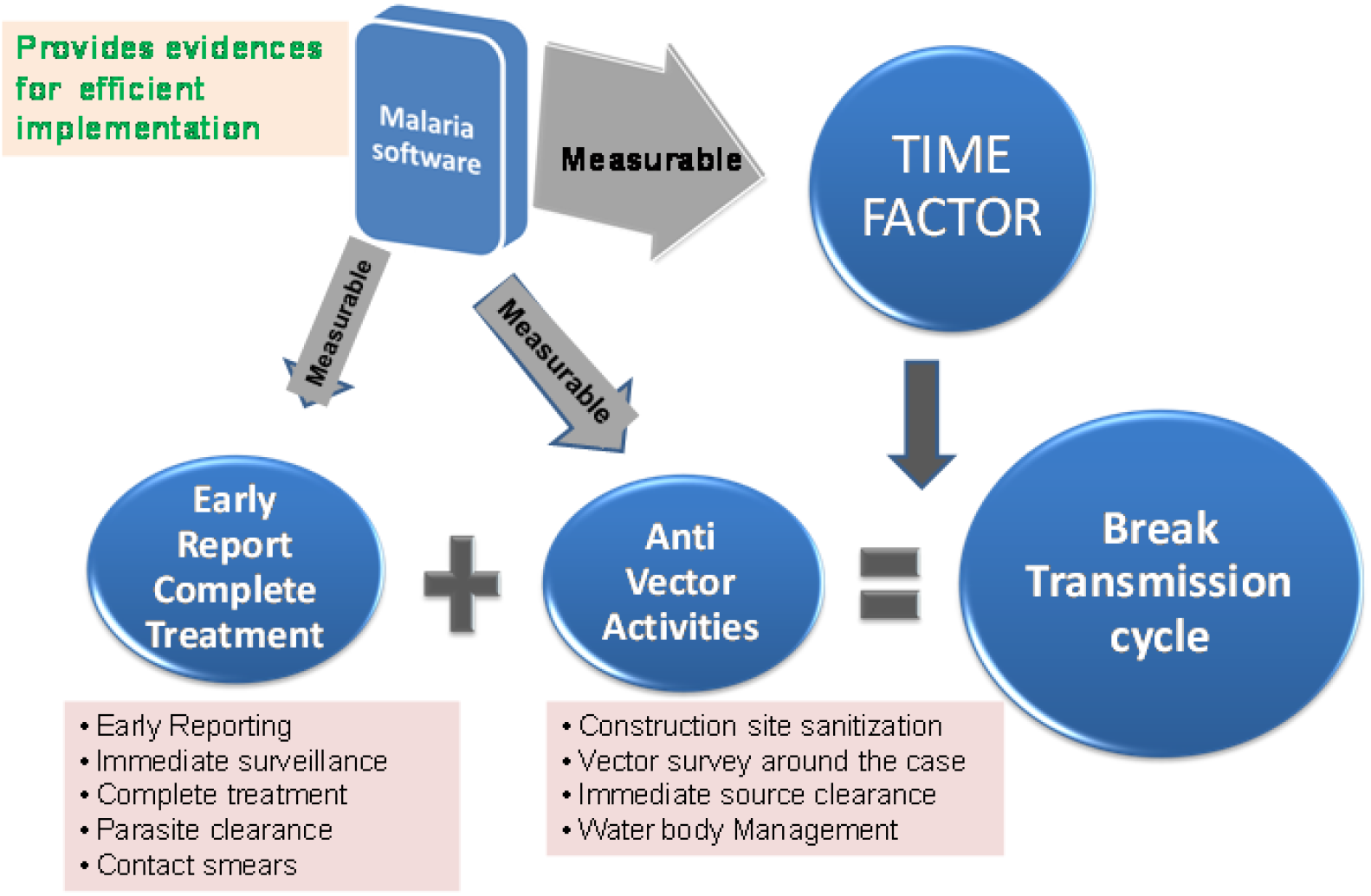
Information technology (IT) logics.

In the 5^th^ year post-digitization, incidence of malaria is further reduced as compared to corresponding period of previous year. Hence, with continued MCS use, other administrative measures and action taken to address issues based on data received via MCS and feedback from stakeholders, the reduction in incidence of malaria was sustained.

## Conclusion

The digital surveillance system coupled with field action and creation of big data has been effective tool to improve systems. Software helped to improve incidence-centric active surveillance, complete treatment with documentation of elimination of parasite, targeted vector control measures. The learnings and analytical output from the data helped to modify strategies for local control of both disease and the vector.

## Data Availability

The data used in this study are archived with Dr BS Baliga and available from them upon reasonable request.

## Abbreviations

TABs: Tablets
GIS: Geographic Information System
NVBDCP: National Vector Borne Disease Control Programme
MPWs: Multi Purpose Workers
MCS: Malaria Control Software
DY: Digitization year
ASARC: Active Surveillance Around Reported Case
ABER: Annual Blood Examination Rate
API: Annual Parasite Incidence
SPR: Slide Positivity Rate
SFR: Slide Falciparum Rate
IEC: Information Education and Communication
IT: Information Technology
MCC: Mengaluru City corporation

## Acknowledgements

Administration of Mangaluru City Corporation and District Health Officials, Dakshina Kannada for accepting and utilizing the software. Assistance from Health workers from the City Corporation and District Malaria Office is acknowledged. Akansha Baliga for assistance in analysis of data, and Dr Chaitali Ghosh for copy editing of the manuscript.

## Funding

No external funding received.

## Authors’ contributions

BSB, NKo and SB conceived the study. BSB and NKo developed the software. AJ, MK and NKo performed all programme implementation. SKG and BGPK for additional technical support. BSB, AJ,SB, and SKG drafted the manuscript. BSB, AJ, NKU and SKG for statistical analysis. All authors read, reviewed and approved the final manuscript

## Ethics declarations

Ethical approval Institutional Ethics Committee, Kasturba Medical College, Mangaluru, India gave opinion as ‘not required’.

## Consent for publication

Not applicable

## Competing interests

The authors declare that they have no competing interests.

## Notes

### Competing Interest Statement

The authors have declared no competing interest.

### Clinical Trial

This is not a clinical trial

### Author Declarations

Institutional Ethics Committee, Kasturba Medical College, Mangaluru, India gave opinion as not required

